# Data Resource Profile: Understanding the patterns and determinants of health in South Asians - South Asia Biobank

**DOI:** 10.1101/2020.08.12.20171322

**Authors:** Peige Song, Ananya Gupta, Ian Y Goon, Mehedi Hasan, Sara Mahmood, Polly Page, Rajendra Pradeepa, Samreen Siddiqui, Wnurinham Silva, Garudam R Aarthi, Saira Afzal, Sophie E Day, Gary S Frost, Bridget A Holmes, Rajan Kamalesh, Dian Kusuma, Marisa Miraldo, Elisa Pineda, Fred Hersch, Baldeesh K Rai, Malabika Sarker, Franco Sassi, Jonathan Valabhji, Nick J Wareham, Sajjad Ahmed, Ranjit M Anjana, Soren Brage, Nita G Forouhi, Sujeet Jha, Anuradhani Kasturiratne, Prasad Katulanda, Khadija I Khawaja, Marie Loh, Malay K Mridha, Ananda R Wickremasinghe, Jaspal S Kooner, John C Chambers

## Abstract

**Background and aims:** This paper describes the data resource profile of South Asia Biobank (SAB), which was set up in South Asia from November 2018 to March 2020, to identify the risk factors and their complex interactions underlying the development of type-2 diabetes mellitus, cardiovascular disease and other chronic diseases in South Asians.

**Data resource basics:** This cross-sectional population-based study has recruited 52713 South Asian adults from 118 surveillance sites at five centres of excellence in South Asia (Bangladesh, North India, South India, Pakistan and Sri Lanka). Structured assessments of participants included six complementary domains: i). Registration and consent; ii). Questionnaire (information on behavioural risk factors, personal and family medical history, medications, socioeconomic status); iii). Physical measurements (height, weight, waist and hip circumference and bio-impedance for body fat composition, blood pressure, cardiac evaluation, retinal photography, respiratory evaluation); iv). Biological samples (blood and urine); v). Physical activity monitoring and vi). Dietary intake by a 24-hour recall. Aliquots of whole blood, serum, plasma, and urine were put into storage at −80°C for further analysis.

**Key results:** The prevalence of obesity is 6.6% in Bangladesh, 19.7% in India, 33.9% in Pakistan and 15.7% in Sri Lanka. The prevalence of diabetes is 11.5%, 27.7%, 25.3%, and 24.8%, and the prevalence of hypertension is 26.7%, 36.9%, 44.5%, 35.0% in Bangladesh, India, Pakistan and Sri Lanka respectively.

**Collaboration and data access:** SAB is the first comprehensive biobank of South Asian individuals. Collected data are available to the global scientific community upon request.

## Data resource basics

Type-2 diabetes mellitus (T2DM) and cardiovascular disease (CVD) are leading and closely interlinked global health challenges ^1^. The burdens of T2DM and CVD are especially high in South Asia, one of the most populous and the most densely populated regions of the world ^3, 4^. The prevalence of diabetes in South Asia has risen more rapidly than in other large geographic regions,^5^ while it is projected that South Asia will account for 40% of the global CVD burden by 2020 ^6^. In addition, T2DM and CVD develop at an earlier age in South Asians than in their European counterparts ^7, 8^.

Identification of the primary risk factors for T2DM and CVD is central to the development of effective approaches for the prevention and treatment of chronic diseases such as T2DM and CVC ^9^. However, epidemiological data are currently sparse for South Asia, with evidence on the drivers of T2DM and CVD being predominantly based on cross-sectional studies that recorded a narrow range of exposures, and without longitudinal assessments^4,6,7,10^. The few available prospective studies are largely derived from investigations of South Asians residing in Western countries, and are further limited by small sample size and incomplete phenotypic characterisation ^4^. To better understand the wide range of exposures that contribute to the development of T2DM and CVD in South Asians, a large-scale population-based study that collects information on demographic, lifestyle, clinical, environmental, and genomic variables is needed.

To address this important need, we have established a unique cross-sectional population study focussed on the South Asian population-South Asia Biobank (SAB). SAB was launched in 2018 as a partnership between collaborating centres in Bangladesh, India, Pakistan, Sri Lanka and the UK ^11^. SAB includes rich baseline demographic, lifestyle, clinical, environmental, and phenotypic data, biological samples from more than 50,000 South Asian participants. This resource will enable a broad range of epidemiological research, including the development of prevention and treatment approaches, discovery of novel molecular biomarkers, risk stratification algorithms and innovative therapeutic approaches for better prevention of T2DM and CVD in South Asians. The specific initial objectives of SAB are to:

1. Establish a network of NCD surveillance sites in Bangladesh, India, Pakistan and Sri Lanka, using common protocols and platforms, in partnership with regional centres of excellence in South Asia;
2. Complete structured health assessments on a representative sample of at least 50,000 South Asians aged 18 years and above residing at all surveillance sites;
3. Use the data to identify the genetic and environmental factors underlying noncommunicable diseases in South Asians, and translate the findings into new approaches for maintenance of health and well-being.

## Data collected

SAB is a cross-sectional population-based study that recruited participants in five study regions: Bangladesh, South India and North India, Pakistan and Sri Lanka. Health information and biological samples of 52713 South Asians were collected from 118 surveillance sites. Recruitment started in November 2018 and ended in March 2020 (due to the pandemic of COVID-19).

### Inclusion and exclusion criteria

We recruited men and women of self-reported South Asian ethnicity aged 18 years and above. We excluded women who were currently pregnant, as well as people who were not permanent residents of the surveillance site (residence for 12 months or more). We also excluded people with serious illness expected to reduce life expectancy to less than 12 months, those who plan to leave the surveillance site within the next 12 months, and those unable or unwilling to give informed consent.

### Study settings and participants

The surveillance sites at which recruitment occurred in each study region are summarised in Supplementary Table 1 (available as Supplementary data at IJE online). Governmental census data and available household listings were used, together with house-house visits by research teams and local primary care workers, to identify (enumerate) the resident population. In each household, demographic details of the eligible adults were obtained, and all people meeting study entry criteria were invited to take part in. We worked closely with community senior members (e.g. teachers, employers, religious leaders) to support and facilitate engagement in the study. Explanations of the project’s purpose were provided in writing and using videos, in relevant South Asian languages, supported by bilingual translators.

**Table 1.**
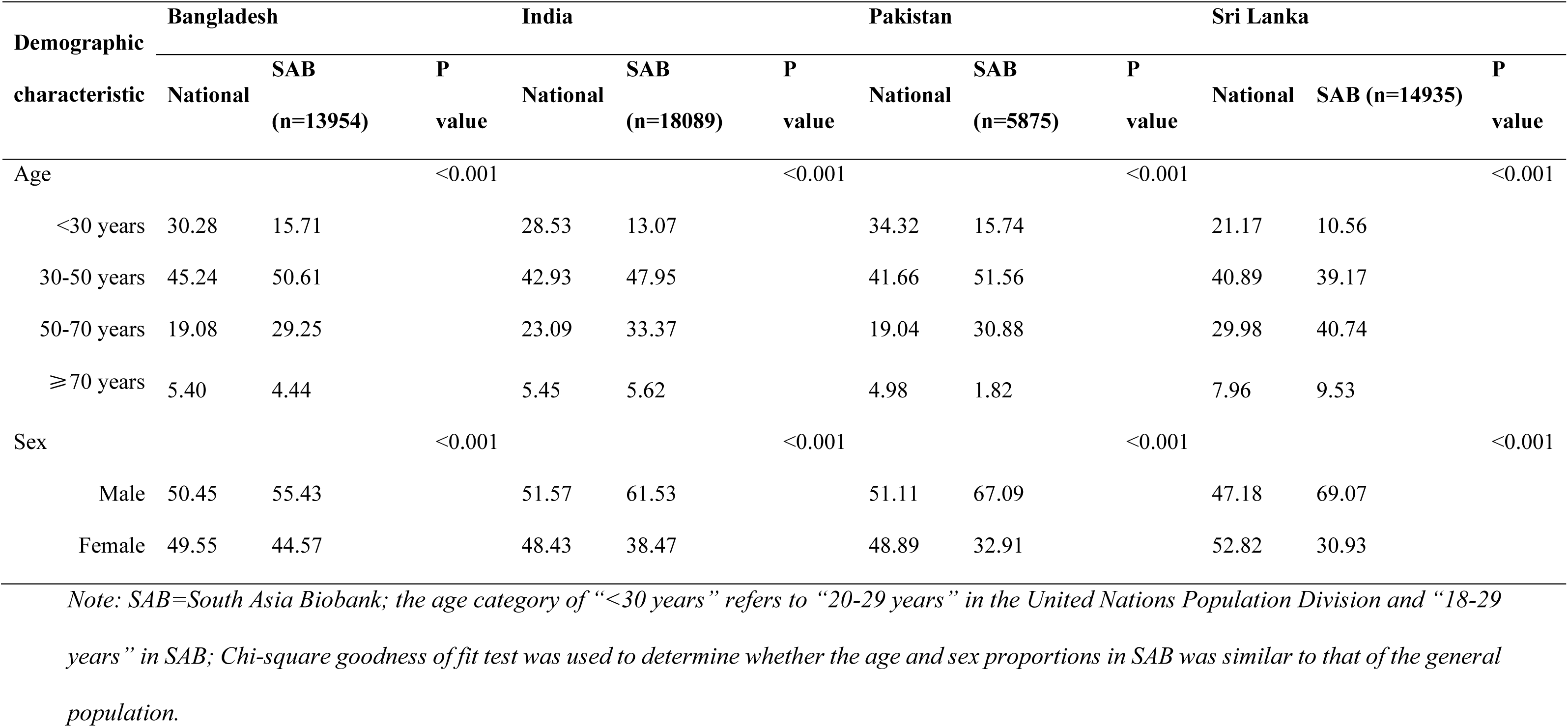
Characteristics of participants in South Asia Biobank (SAB), compared to reported national population distribution (United Nations Population Division)

| Demographic characteristic | Bangladesh |  |  | India |  |  | Pakistan |  |  | Sri Lanka |  |  |
| --- | --- | --- | --- | --- | --- | --- | --- | --- | --- | --- | --- | --- |
|  | National | SAB (n=13954) | P value | National | SAB (n=18089) | P value | National | SAB (n=5875) | P value | National | SAB (n=14935) | P value |
| Age |  |  | <0.001 |  |  | <0.001 |  |  | <0.001 |  |  | <0.001 |
| <30 years | 30.28 | 15.71 |  | 28.53 | 13.07 |  | 34.32 | 15.74 |  | 21.17 | 10.56 |  |
| 30-50 years | 45.24 | 50.61 |  | 42.93 | 47.95 |  | 41.66 | 51.56 |  | 40.89 | 39.17 |  |
| 50-70 years | 19.08 | 29.25 |  | 23.09 | 33.37 |  | 19.04 | 30.88 |  | 29.98 | 40.74 |  |
| ≥70 years | 5.40 | 4.44 |  | 5.45 | 5.62 |  | 4.98 | 1.82 |  | 7.96 | 9.53 |  |
| Sex |  |  | <0.001 |  |  | <0.001 |  |  | <0.001 |  |  | <0.001 |
| Male | 50.45 | 55.43 |  | 51.57 | 61.53 |  | 51.11 | 67.09 |  | 47.18 | 69.07 |  |
| Female | 49.55 | 44.57 |  | 48.43 | 38.47 |  | 48.89 | 32.91 |  | 52.82 | 30.93 |  |

*Note: SAB=South Asia Biobank; the age category of “<30 years” refers to “20-29 years” in the United Nations Population Division and “18-29 years” in SAB; Chi-square goodness of fit test was used to determine whether the age and sex proportions in SAB was similar to that of the general population.*

By March 2020 we recruited a total of 52853 subjects: 13954 from Bangladesh, 8620 from South India, 9469 from North India, 5875 from Pakistan and 14935 from Sri Lanka.

### Measures

Participants were invited to attend the survey sites between 7am and 11am in the fasting state (water only after midnight). Structured assessments of participants were conducted in six complementary domains: i. Registration and consent; ii. Health and lifestyle questionnaire; iii. Physical measurements; iv. Biological samples (blood and spot urine); v. Physical activity monitoring and vi. 24-hour dietary recall. Procedures and training were standardised between countries and surveillance sites.

1. <u>Registration and consent</u>. Written, informed consent was obtained from all participants for data collection, and inclusion in the research. Informed consent included permission for the data and samples collected to be used for chronic disease research, including data sharing with national and international bodies concerned with prevention and control of T2DM and CVD, as well as for molecular epidemiological research. Consent was facilitated using videos (available in major South Asian languages). A unique study ID was allocated to each participant.
2. <u>Questionnaire</u>. An interviewer-administered health and lifestyle questionnaire was used to collect information on behavioural risk factors (smoking, alcohol use, physical activity and consumption of fruits/vegetables), personal and family medical history, medications, and socio-economic status. The questionnaire was founded on the extended WHO STEPwise approach to Surveillance (STEPS) questionnaire that is widely used in global disease surveillance, which was adapted for use in South Asia context, through incorporating additional questions ^12^.
3. <u>Physical measurements</u>. These included: a) Anthropometry (height, weight, waist and hip circumference and bio-impedance for body fat composition); b) Blood pressure by digital device; c) Cardiac evaluation by 12 lead ECG to identify arrhythmia, left ventricular hypertrophy and previous myocardial infarction; d) Retinal photography for assessment of retinal disease, including hypertensive and diabetic retinopathy; and e). Respiratory evaluation by spirometry to assess for smoking/environment-related lung injury.
4. <u>Biological samples</u>. 25ml venous blood was collected using venesection by trained phlebotomists and then distributed into EDTA, serum and citrate vacutainer tubes, and into tubes designed for RNA preservation (Tempus tube). Fasting glucose, and cholesterol were measured by point of care tests. An Oral Glucose Tolerance Test was carried out in a subset of participants, enabling validation of diabetes classification. A spot urine sample (6 ml in 3 aliquots) was also collected for analysis of albuminuria and other biomarkers. Aliquots of whole blood, buffy coat, serum, EDTA plasma, citrate plasma, and urine (Supplementary Table 2, available as Supplementary data at IJE online) were stored at −80℃ for future molecular epidemiological research (including genomics) to investigate the mechanisms underpinning the development of T2DM and CVD, and other complex diseases that are of importance to South Asians (including but not limited to: obesity, cancer, dementia, COPD, chronic kidney disease).
5. <u>Physical activity</u> was also objectively quantified in 100Hz resolution using a wrist-worn triaxial accelerometer, worn on participants’ non-dominant wrist for seven days. This device is small, light-weight, wrist-watch shaped, battery-powered and uses triaxial acceleration in gravitational units to infer participant movement. It has been used recently to measure physical activity patterns amongst 100,000 people in the UK Biobank study ^13^.
6. <u>Dietary</u> intake was recorded by interviewer-administered computerised 24-hour dietary recall based on the multiple pass method using the Intake24 system (https://intake24.org/). The system was specifically adapted for the South Asian context through incorporating extensive additional foods, drinks and dishes, and portion size photograph relevant to the study settings. Adaptation was informed by research nutritionists and dieticians from each study centre and by the results of previous dietary surveys in the study locations. The implementation of this tool could enable the description of food and nutrient intakes, evaluation of intakes in comparison with guidelines, and the investigation of the link between diet and health endpoints.

All study participants received a report summarising the clinically relevant results of their health assessment, together with an explanatory booklet and a link to access an explanatory video. Participants identified with significant health conditions (e.g. T2DM, hypertension) had the opportunity to discuss the results with the study team, and to be referred to an appropriate healthcare facility for further assessment, counselling or treatment.

### Environmental mapping

In each surveillance site, an environmental mapping exercise was carried out. The aim was to characterise the built environment in terms of retailers and advertisements for food and tobacco and physical activity facilities. The methodologies were adapted from food modules conducted by the International Network for Food and Obesity/NCDs Research, Monitoring and Action Support (INFORMAS), the Maryland Food Systems Map conducted by the Johns Hopkins Center for a Livable Future, and the World Health Organization Framework Convention on Tobacco Control ^14-16^. In addition to geolocations, the main variables included food (e.g., fruit, vegetables, confectionery), drinks (e.g., soft drinks, sugar-free drinks), and tobacco products (e.g., cigarette, beedi) being sold or advertised. Data collection used KoboToolBox for Android (https://www.kobotoolbox.org) and covered each surveillance site with a 500-meter buffer beyond the site boundary.

### Identification of outcomes

The primary outcomes were T2DM and cardiovascular disease. The secondary endpoints included respiratory and chronic kidney diseases, or cancer.

### Quality control and data management

The surveillance teams, comprising research assistants, laboratory technicians, physicians and coordinators, were trained to follow standardised protocols (Supplementary Table 3, available as Supplementary data at IJE online) Their training modules included interviewing techniques, ethics and specific instructions for data variables (demographic, socio-economic, food security, behavioural risk factors, medication and lifestyle practices, physical measurement and collection of biological samples).

Revalidation of the research teams in study procedures was done at regular intervals during the study to ensure high-quality data collection that was harmonised across surveillance sites. Standardised operating procedures were established for all data collection procedures. Questionnaires were translated into the local languages local to the communities, and back-translated. Equipment used for physical and biological measurements is listed in Supplementary Table 4, available as Supplementary data at IJE online, and was regularly calibrated using appropriate controls/standards.

The data management teams reviewed the data collected routinely for completeness and data quality, including using custom computer scripts to assess for biases in data entry, logical inconsistencies, internal correlations, digit preference, measurement drift or bias between machines and observers. Quality control reports were circulated at weekly intervals between the study investigators, to drive continuous evaluation and improvement in study processes. A random subset comprising up to 2% of the study participants, and/or a subset of biological samples were reassessed to provide additional quality control information. Data collection methods used were “field-friendly”, culturally-acceptable and minimally-invasive in order to reduce participant attrition and improve logistical feasibility.

Personal and clinical data were separated by pseudonymisation to enhance data security. All data were encrypted during transmission and stored securely both locally and in a cloud-based infrastructure. Data and all relevant documents will be stored for a minimum of ten years. Samples collected were split and stored in both South Asia and the UK to ensure long-term (>20 years) sample integrity and preservation. While some laboratory assays on stored samples were done in South Asia, the majority of assays were carried out in the UK or other countries with relevant technologies in the future.

## Data resource use

Data collected in this cross-sectional investigation could be used to assess the epidemiology of T2DM, CVD and other chronic diseases in South Asia. The exploration of possible risk factors for T2DM and CVD could provide a scientific basis for evidence-based public health policymaking and interventions. Upon request, the rich resources of SAB are available to researchers from all over the world.

## Key results and findings

A total of 52853 participants from the four participant countries took part in SAB; the locations of all surveillance sites are demonstrated in Figure 1. The response rate based on enumerated population in each surveillance site ranged from 17.6% in North India to 72.3 % in Pakistan (see Supplementary Table 5 for more details, available as Supplementary data at IJE online). The demographic structure of the study participants and the comparison with the National Population data in 2015, obtained from the United Nations Population Division, are shown in Table 1 ^17^. Based on the definition of obesity by WHO, the prevalence of obesity (body mass index ≥30 kg/m^2^) is 6.6% in Bangladesh, 19.7% in India, 33.9% in Pakistan and 15.7% in Sri Lanka. The prevalence of diabetes, defined as a fasting glucose level >126 mg/dL or a physician-diagnosis, or currently on antidiabetic medications, is 11.5%, 27.7%, 25.3%, and 24.8%, and the prevalence of hypertension, defined as a systolic blood pressure ≥ 140 mmHg, or a diastolic blood pressure ≥ 90 mmHg, or a physician-diagnosis, or currently being on antihypertensive medications, is 26.7%, 36.9%, 44.5%, 35.0% in Bangladesh, India, Pakistan and Sri Lanka respectively.

**Figure 1.**
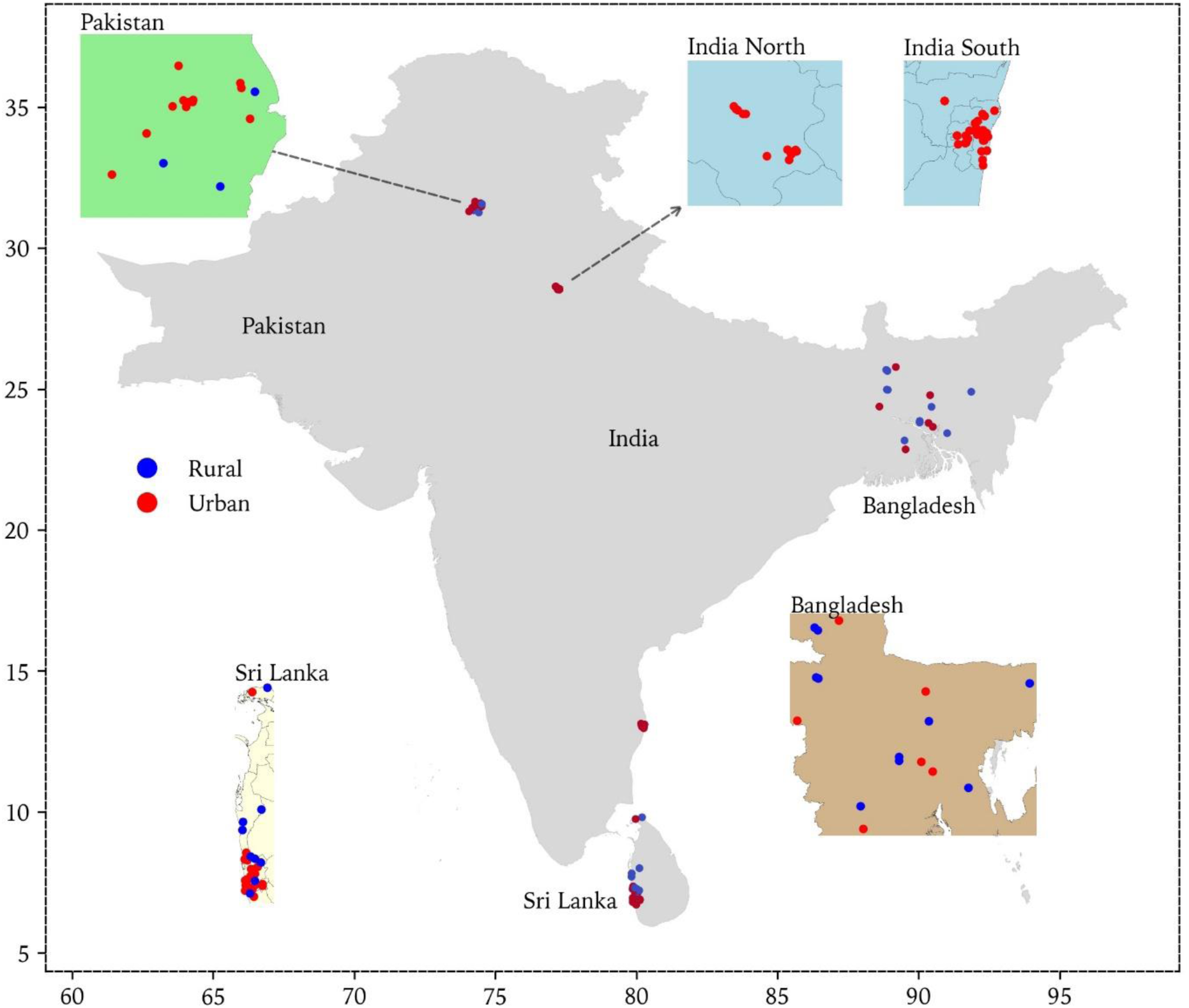
Locations of the South Asia Biobank (SAB) surveillance sites

## Strengths and weaknesses

SAB is designed to identify the risk factors and their complex interactions underlying the development of T2DM, CVD and other chronic diseases in South Asians. With intensive data collection, SAB provides representative population samples in four South Asian countries. SAB is the first comprehensive biobank of South Asian individuals. Its large sample size, broad geographical reach and wide range of data collected, including bio-samples, make SAB a powerful tool for epidemiological and translational research in South Asian populations. The standardised procedures and rigorous quality control of data collection ensure comparability of study results between and within the partner countries. Further, although random sampling approaches were used in selecting participants, we cannot exclude ‘healthy volunteer effects, a common phenomenon in epidemiological research. In addition, advanced phenotyping by imaging (e.g. MRI, DXA or ultrasound) was not feasible across the range of sites studied.

## Data resource access

Reports and major results of SAB will be released regularly on the SAB website (https://www.ghru-southasia.org/). Subject to data privacy requirements, and the permissions included in the consent form, individual-level data and samples are available for use to approved investigators.

## Supplementary Data

Supplementary data are available at IJE online.

## SAB in a nutshell

The South Asia Biobank (SAB) is a large, cross-sectional population-based study of the risk factors and their interactions in the development of T2DM, CVD and other chronic diseases in South Asians. Recruitment, starting in November 2018 and ending in March 2020, was through all surveillance sites at five centres of excellence in South Asia (Bangladesh, North India, South India, Pakistan and Sri Lanka). The study aimed to recruit more than 50,000 adults in South Asia by March 2020. Structured assessments of participants included six complementary domains: i). Registration and consent; ii). Questionnaire (information on behavioural risk factors, personal and family medical history, medications, socioeconomic status); iii). Physical measurements (height, weight, waist and hip circumference and bio-impedance for body fat composition, blood pressure, cardiac evaluation, retinal photography, respiratory evaluation); iv). Biological samples (blood and urine); v). Physical activity monitoring and vi). Dietary intake by a 24-hour recall. Aliquots of whole blood, serum, plasma, and urine were put into storage at −80°C for further analysis. Collected data are available to the global scientific community upon request. Reports and major results of SAB will be regularly released. Potential collaborative research is invited.

## Ethics

SAB was conducted in accordance with the recommendations for physicians involved in research on human subjects adopted by the 18th World Medical Assembly, Helsinki 1964 and later revisions. Research approval was obtained from the Imperial College London Research Ethics Committee (reference: 18IC4698) and local Institutional Review Boards in each of the participating countries.

## Data Availability

Reports and major results of SAB will be released regularly on the SAB website. Subject to data privacy requirements, and the permissions included in the consent form, individual-level data and samples are available for use to approved investigators.

https://www.ghru-southasia.org/

## Acknowledgements

All authors thank all the team members and all participants in the South Asia Biobank. Conflict of interest: None declared.

## References

1. Roth GA, Johnson C, Abajobir A, et al. Global, regional, and national burden of cardiovascular diseases for 10 causes, 1990 to 2015. Journal of the American College of Cardiology 2017; 70: 1–25.

2. Ogurtsova K, da Rocha Fernandes J, Huang Y, et al. IDF Diabetes Atlas: Global estimates for the prevalence of diabetes for 2015 and 2040. Diabetes research and clinical practice 2017; 128: 40–50.

3. Misra A, Tandon N, Ebrahim S, et al. Diabetes, cardiovascular disease, and chronic kidney disease in South Asia: current status and future directions. bmj 2017; 357: j1420.

4. Dans A, Ng N, Varghese C, Tai ES, Firestone R, Bonita R. The rise of chronic noncommunicable diseases in southeast Asia: time for action. The Lancet 2011; 377: 680–9.

5. Ghaffar A, Reddy KS, Singhi M. Burden of noncommunicable diseases in South Asia. Bmj 2004; 328: 807–10.

6. Gholap N, Davies M, Patel K, Sattar N, Khunti K. Type 2 diabetes and cardiovascular disease in South Asians. Primary Care Diabetes 2011; 5: 45–56.

7. Forouhi N, Merrick D, Goyder E, et al. Diabetes prevalence in England, 2001—estimates from an epidemiological model. Diabetic Medicine 2006; 23: 189–97.

8. Forouhi N, Sattar N, Tillin T, McKeigue P, Chaturvedi N. Do known risk factors explain the higher coronary heart disease mortality in South Asian compared with European men? Prospective follow-up of the Southall and Brent studies, UK. Diabetologia 2006; 49: 2580–8.

9. Eckel RH, Kahn R, Robertson RM, Rizza RA. Preventing cardiovascular disease and diabetes: a call to action from the American Diabetes Association and the American Heart Association. Circulation 2006; 113: 2943–6.

10. Organisation WH. Noncommunicable diseases in the South-East Asia Region, 2011: situation and response. 2012.

11. Sudlow C, Gallacher J, Allen N, et al. UK biobank: an open access resource for identifying the causes of a wide range of complex diseases of middle and old age. PLoS medicine 2015; 12: e1001779.

12. Organisation WH. WHO STEPS surveillance manual: the WHO STEPwise approach to chronic disease risk factor surveillance: Geneva: World Health Organization; 2005.

13. Doherty A, Jackson D, Hammerla N, et al. Large scale population assessment of physical activity using wrist worn accelerometers: the UK biobank study. PloS one 2017; 12: e0169649.

14. INFORMAS. International Network for Food and Obesity/noncommunicable Disease Research, Monitoring and Action Support. [cited July 20, 2018]; Available from: http://www.informas.org/

15. Misiaszek C, Buzogany S, Freishtat H. Baltimore City’s food environment: 2018 report. Johns Hopkins Center for a Livable Future (January 2018) Accessed April 2018; 19.

16. Organisation WH. Report on the Global Tobacco Epidemic. The MPOWER package. Geneva: World Health Organization 2008. 2008.

17. United Nations DoE, Social Affairs PD. World Population Prospects 2019. United Nations New York; 2019.

